# Clinical Gait Analysis for Hip Osteoarthritis Diagnostic Model and Arthroplasty Treatment’s Evaluation

**DOI:** 10.1101/2023.04.28.23289220

**Authors:** Weiheng Zhang

## Abstract

Osteoarthritis (OA) is a degenerative joint disease that seriously disturbs the patients’ motor ability [7]. To quantify gait kinematic impairments from such bone diseases, clinical gait analysis is widely used. Based on the motion-captured gait analysis data published by Bertaux et al., stratified by the subjects’ body mass index (BMI), we built a computational pipeline to extract seven gait kinematic features that can significantly distinguish hip OA (HOA) patients from healthy subjects [3], [5]. These features, along with demographic variables, were utilized to fit regression models for HOA predictor interpretation and classification models for HOA diagnosis. CV results showed that the final diagnostic model could achieve 90%+ sensitivity under 90% specificity, with AUC up to 0.96. We also analyzed the HOA patients 6 months post arthroplasty treatment: patients showed a trend of improvement in most kinematics variables and Ground Reaction Force similarities compared to healthy controls. Although differences between the treated patients and the healthy control still persist. Clinical gait analysis, along with data science strategies, can have substantial potential for HOA diagnosis and evaluation of the arthroplasty treatment.

## II. Introduction

Osteoarthritis (OA), the most common type of arthritis, is a degenerative joint disease that breaks down the joint tissue over time [7]. According to the Centers for Disease Control and Prevention, OA is affecting more than 32.5 million adults in the United States in the year 2023 [7]. OA on the hip joints would result in significant pain and seriously disturb the patient’s motor ability. Patients with end-stage hip OA are usually treated with total hip arthroplasty (THA), which is a cost-effective surgery to relieve pain and restore joint function [2], [3].

In occupational therapy, clinical gait measurements are widely used to quantify gait impairments caused by bone diseases. Dr. Bertaux’s team from Univ. of Bourgogne Franche-Comté conducted such measurements on a total of 80 healthy volunteers and 106 unilateral hip OA (HOA) patients [3]. Each subject walked along a 6-meter straight line with force-sensing plates and motion capture sensors recording their body motion and step forces [3]. Some of the patients 6 months after total hip arthroplasty participated in such measurements again. [3]. The dataset of this clinical gait measurement was publicly available on Nature Scientific Data [3].

Clinical gait measurements show promise for assessing gait deviations and impairments in HOA patients, but the consensus is lacking on which kinematic features to use for diagnosis and follow-up [3]. Based on Bertaux et al.’s dataset, stratified by the subjects’ body mass index (BMI), we built a computational pipeline to extract seven gait kinematic features that can significantly distinguish HOA patients from healthy subjects. We utilized these features, along with demographic variables, to fit regression models for HOA predictor interpretation and classification models for HOA diagnosis. CV results showed that the final diagnostic model could achieve 90%+ sensitivity under 90% specificity, with AUC up to 0.96. We also analyzed the patients 6 months post-treatment: patients showed a trend of improvement in most kinematics variables and Ground Reaction Force similarities compared to healthy controls. Although differences between the treated patients and the healthy control still persist. Clinical gait analysis, along with data science strategies, can have substantial potential for HOA diagnosis and evaluation of the arthroplasty treatment.

## III. Methods

### Study Group

Dr. Bertaux’s team recruited 80 healthy volunteers and 108 end-stage unilateral hip OA patients in the Dijon University Hospital of France, between 2011 and 2016 [3]. The team recorded each subject’s demographic and body figure data. Each subject was asked to wear a motion capture suit and walked naturally back and forth along a 6-meter straight walkway with two embedded force plates [3]. To avoid conscious adaptation, the subjects were not informed about the specific location of the force plates [3]. Ideally, a healthy subject should have one foot stepping on the first plate, followed by another foot stepping on the second plate. Stepping on the connection between two plates would not generate usable ground reaction force. HOA patients with shorter step lengths may only align one foot with the force plate in one trial, and switch foot in the next trial. A minimum of 10 walking trials were performed for each subject. In our analysis, we selected the trial with clear and separate steps on each plate.

Elderly and obesity have been found to significantly affect walking kinematics [5]. Specifically, Fang et al. showed that subjects aged 70 or older had significantly lower walking speed and cadence compared with all other age groups [5]. Also, even after normalizing according to weights, normal-weight (18 <= BMI < 25), overweight (25 <= BMI < 30), and obese (BMI > 30) adults were found to have significantly different walking speed and cadence [5]. Based on these previous results, our project excluded all subjects aged 70 or older. We also stratified the subjects into normal-weighted, overweight, and obese groups using the same BMI criteria. Only subjects in the same weight group would be statistically compared.

### General Gait Kinematics Features

We coded computational pipelines in R 4.1.1 to extract the following gait kinematic features from the data captured by the motion capture system [1], [3], [5]:

♦ Speed (mm/s): Average moving speed of the subject’s center-of-mass (COM). The COM was automatically generated by the Vicon Nexus motion capture software.
♦ Step length (mm): The average distance between the point of initial contact of one foot and the point of initial contact of the opposite foot.
♦ Step length difference (mm): Right and left step lengths are expected to be similar for healthy subjects but may not apply to HOA patients. Thus, we calculated the average difference between the left and right step lengths for each subject.
♦ Cadence (step): Number of steps per minute.
♦ Hold time (s): Average time between foot contact to foot leaving the floor.
♦ Hold time difference (s): Similar to step length, right and left foot’s hold times are expected to be similar for healthy subjects, but may not apply for HOA patients. We calculated the average difference between left and right hold time for each subject.

In order to minimize the effect of the subjects’ body figures on the kinematic features, the following normalization of the above features was applied [5]. In our analysis, the mean body height was calculated separately for each weight group.

♦ norm speed (mm/s) = speed / sqrt (body height/mean body height)
♦ norm step length (mm) = step length / (body height/mean body height)
♦ norm step length diff (mm) = | normalized left step length – normalized right step length |
♦ norm cadence (step) = cadence × sqrt (body height/mean body height)
♦ norm hold time (s) = hold time / sqrt (body height/mean body height)
♦ norm hold time difference (s) = | normalized left hold time – normalized right hold time |

### Ground Reaction Force Analysis

The two force plates collected the ground reaction force (GRF) with a sampling frequency of 1000 Hz [3]. The raw forces were normalized by Vicon Nexus software’s internal algorithm using the subject’s weight [3].

From our exploratory data analysis, a healthy subject’s GRF curve usually has two high peaks, with a deep smooth concave between them. While a HOA patient’s GRF curve tends to have a longer horizontal span, lower peaks, and a shallower, less smooth concave in between. Previous literature defines the first peak as the weight acceptance force, and the second peak as the foot push-off force [8]. The concave between two force peaks represents the mid-stance of a step [8]. We observed that a healthy subject’s weight acceptance and foot push-off peaks tend to be more significant compared to peaks generated by a HOA patient. To quantify the above differences, we propose a succinct and highly effective feature: Area Above Curve (AAC). First, we define a local maximum as the highest point among at least 10 points on both sides. Then, as shown in Figure 1 above, we connect the two local maximums in a GRF curve, resulting in a maxima border. The AAC is then calculated as:

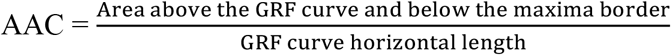

For healthy subjects, the first two local maximum points would be the weight acceptance force peak and the foot push-off force peak. While for HOA patients with fluctuated concave, the second local maximum might come earlier than the foot push-off peak, making the final AAC even more significantly smaller than that from a healthy subject.

**Figure 1.**
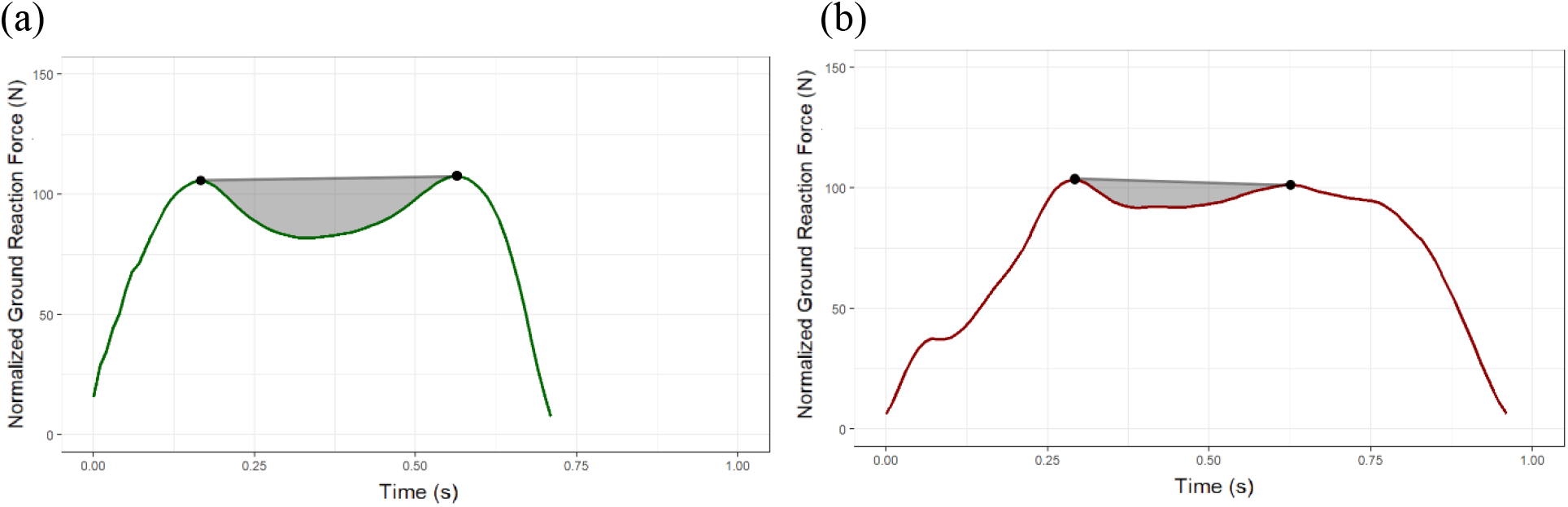
Typical ground reaction force (GRF) curve, along with the Area Above Curve (AAC) shaded in grey generated by: (a). a healthy volunteer. (b) a HOA patient.

**Figure 2.**
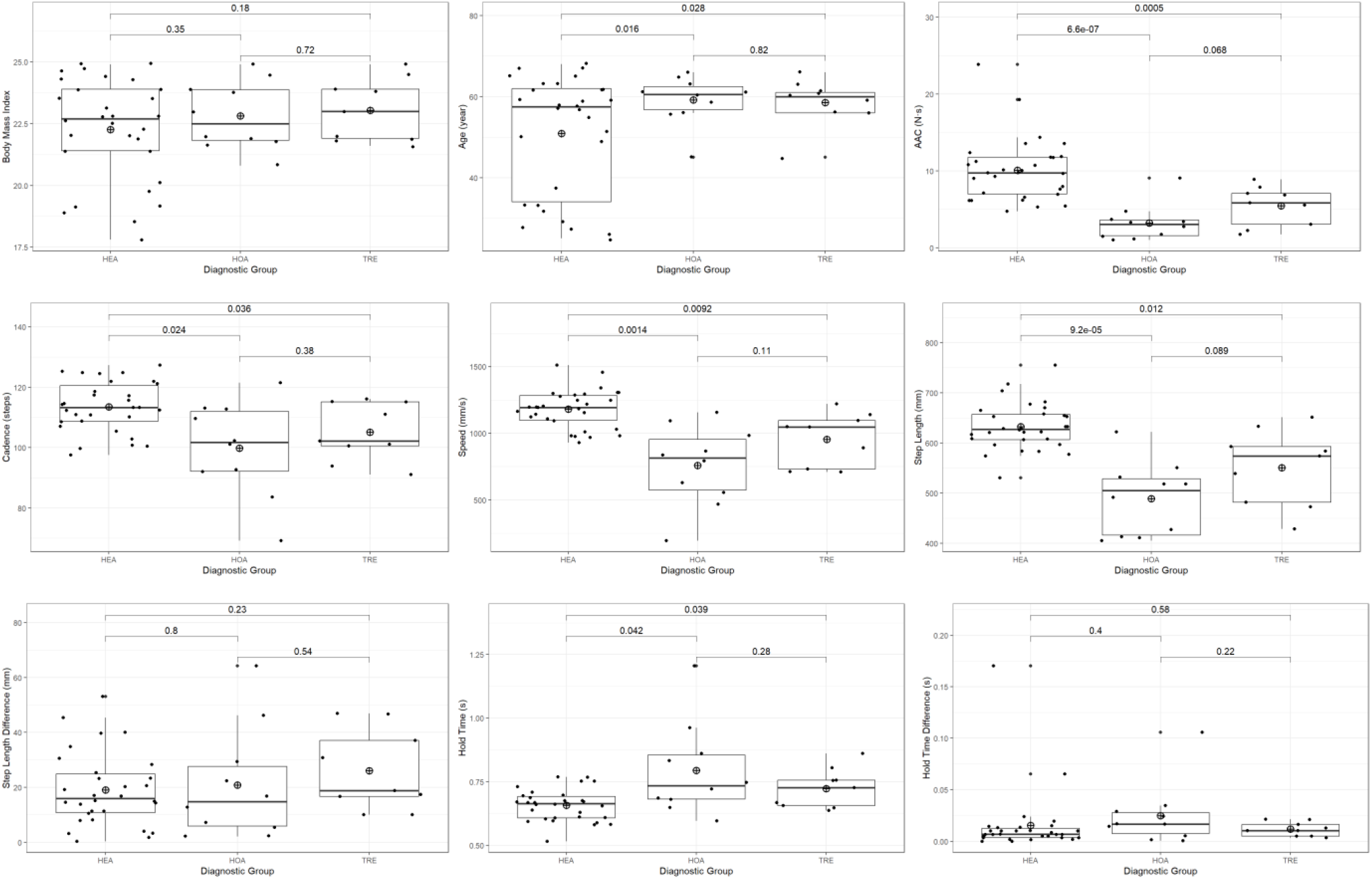
Gait kinematic features among normal weighted subjects. P-values of two-sample independent t-tests between diagnostic groups are labeled on the brackets. The horizontal dashes in the middle of the boxes were medians. The “⨁” markers were group means.

**Figure 3.**
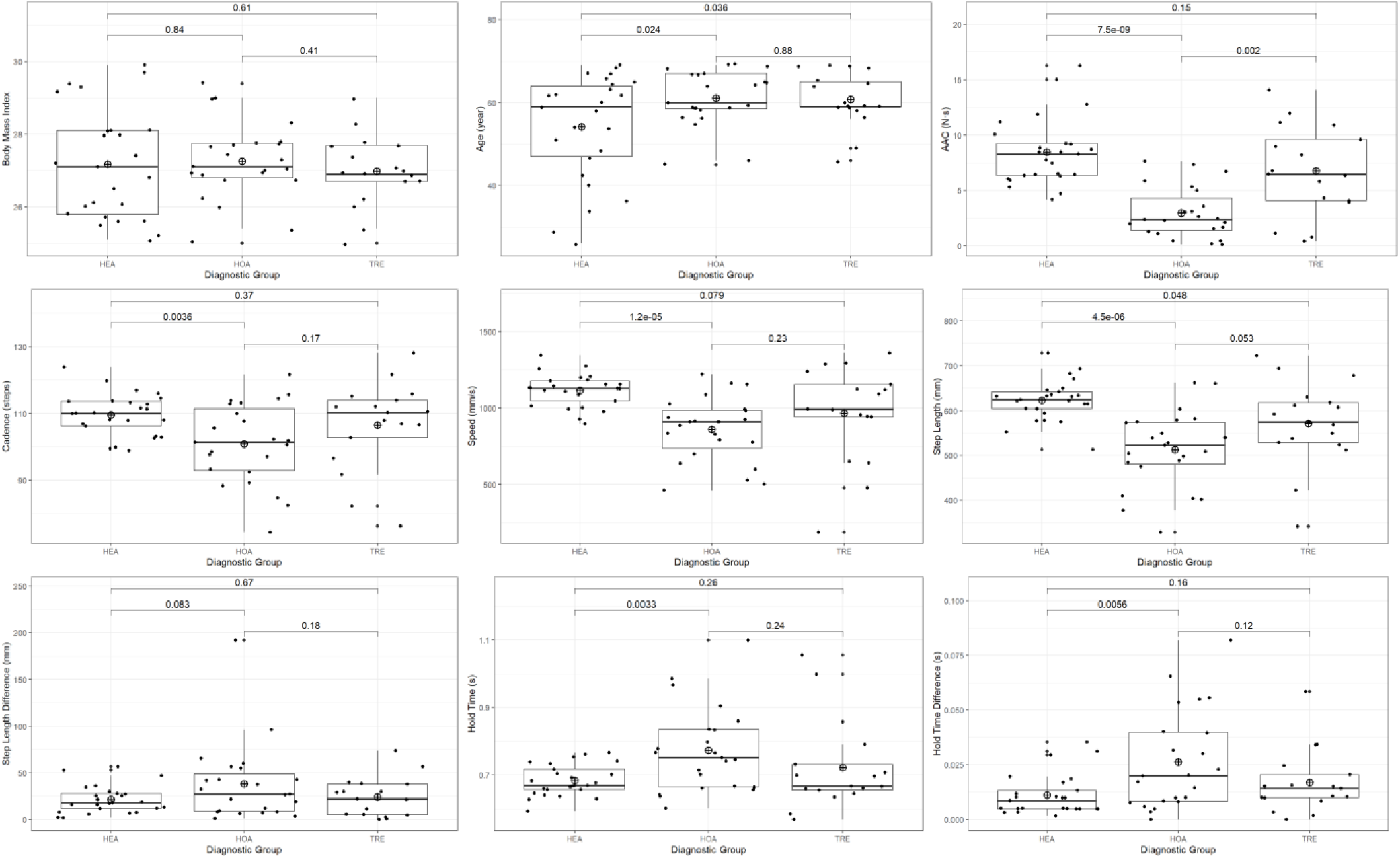
Gait kinematic features among overweighted subjects. P-values of two-sample independent t-tests between diagnostic groups are labeled on the brackets. The horizontal dashes in the middle of the boxes were medians. The “⨁” markers were group means.

**Figure 4.**
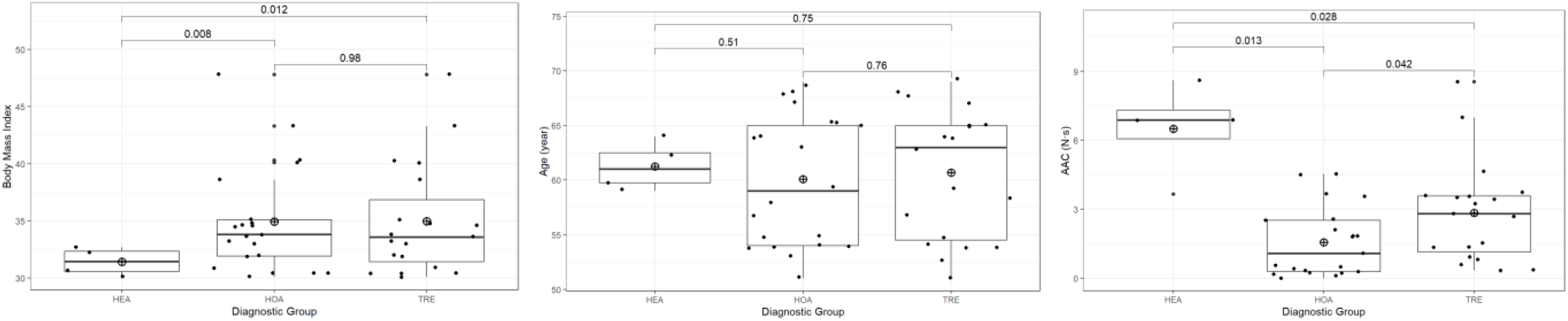
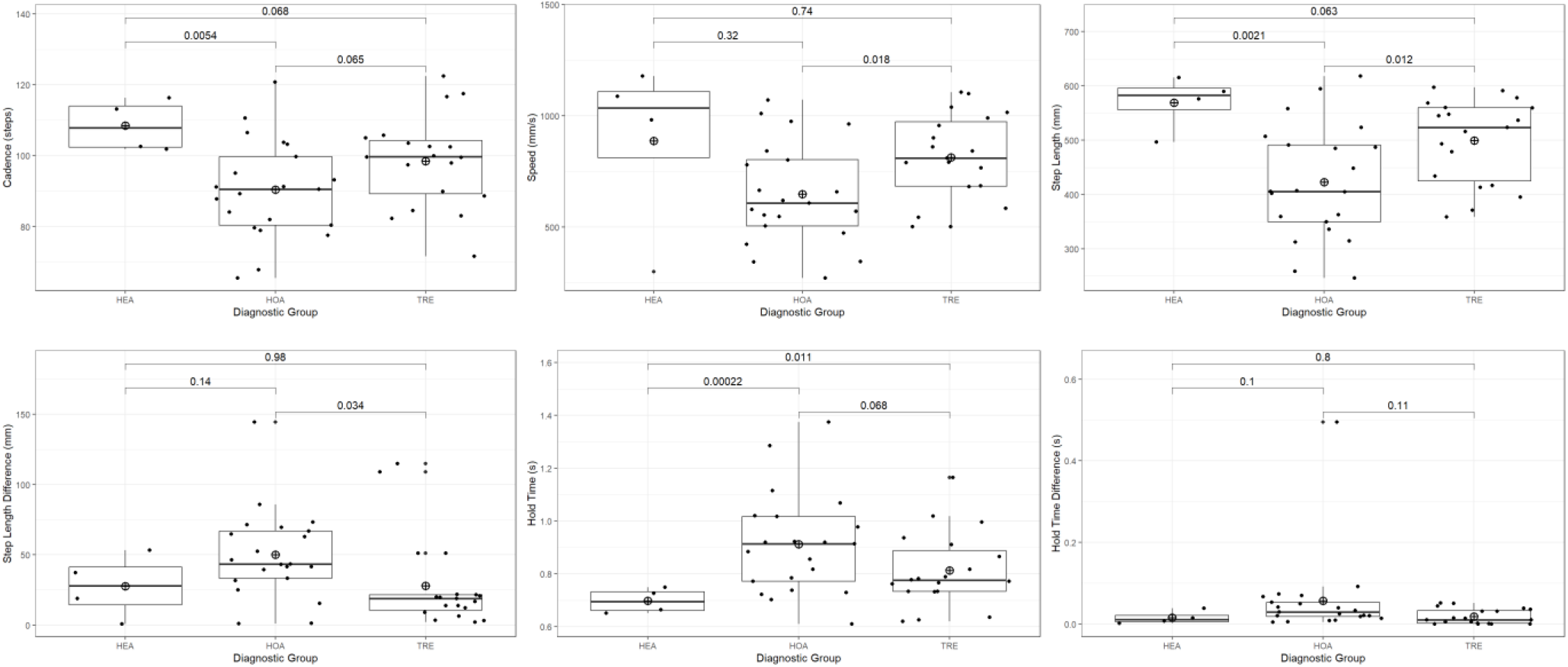
Gait kinematic features among obese subjects. P-values of two-sample independent t-tests between diagnostic groups are labeled in the brackets. The horizontal dashes in the middle of the boxes were medians. The “⨁” markers were group means.

Our further EDA showed that a HOA patient might have a fluctuated GRF on either the HOA-affected foot or the opposite foot. Thus, for each subject, we extracted the GRF curves and calculated AACs for both feet, then select the GRF with a smaller AAC value for further analysis.

### Fitting Penalized Logistic Regression

We used the “caret” package in R 4.1.1 to fit penalized logistic regression for binary classification of healthy volunteers and HOA patients. The model was fitted under 5-fold 3-time repeated cross-validation, using method = “glmnet” and metric = “ROC”. The alpha and lambda of the regression were selected using automatic parameter tuning of the expand.grid() function. Independent models were fitted for the normal-weight subjects, overweight subjects, and all subjects (normal + overweight + obese). The obese group did not have an independent model because only four healthy subjects were available. The model performances were evaluated using “MLeval” package’s evalm() function, generating overall ROC curves and AUC values from cross-validation results.

## IV. Results

### Comparing Features Across Weight Groups and Diagnostic Groups

Table 1 below is the detailed statistics of all variables across all subjects, stratified by weight group and diagnostic group. Same to the results reported by Fang et al., for healthy volunteers (HEA), subjects in a higher weight group tend to walk with a decreased cadence, speed, step length, along with increased step length difference and hold time. AAC, our proposed new feature, also drops significantly as the weight group increases in the HEA group.

**Table 1.**
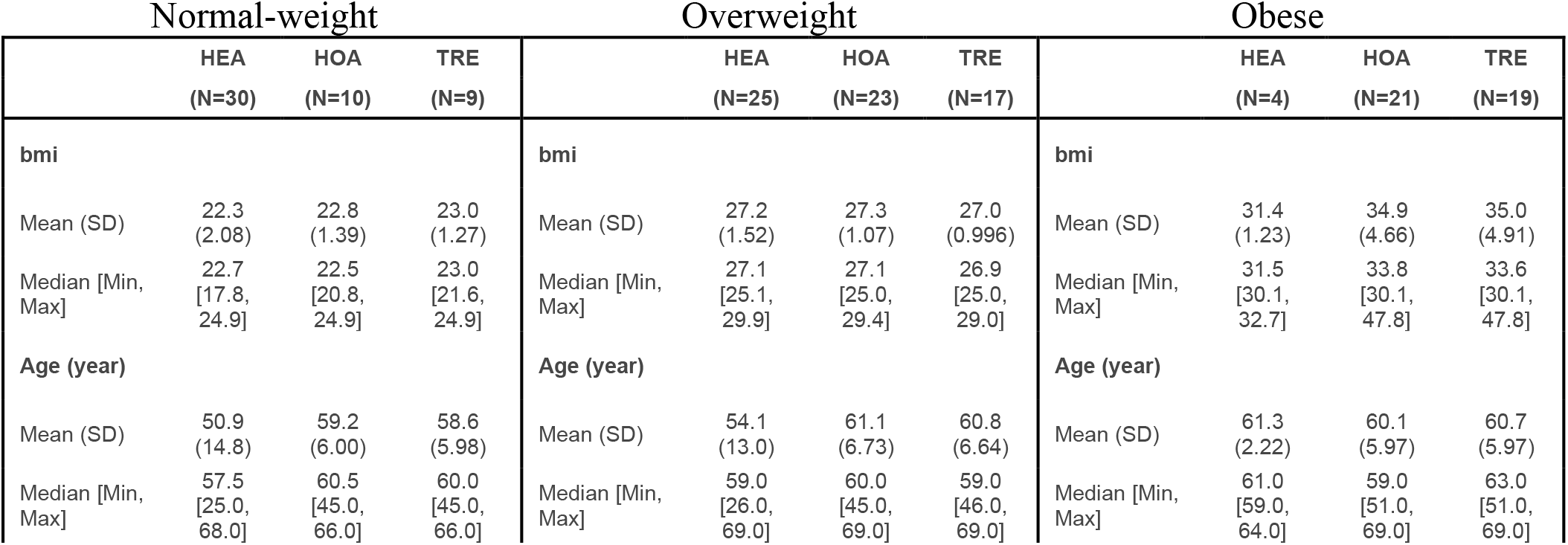
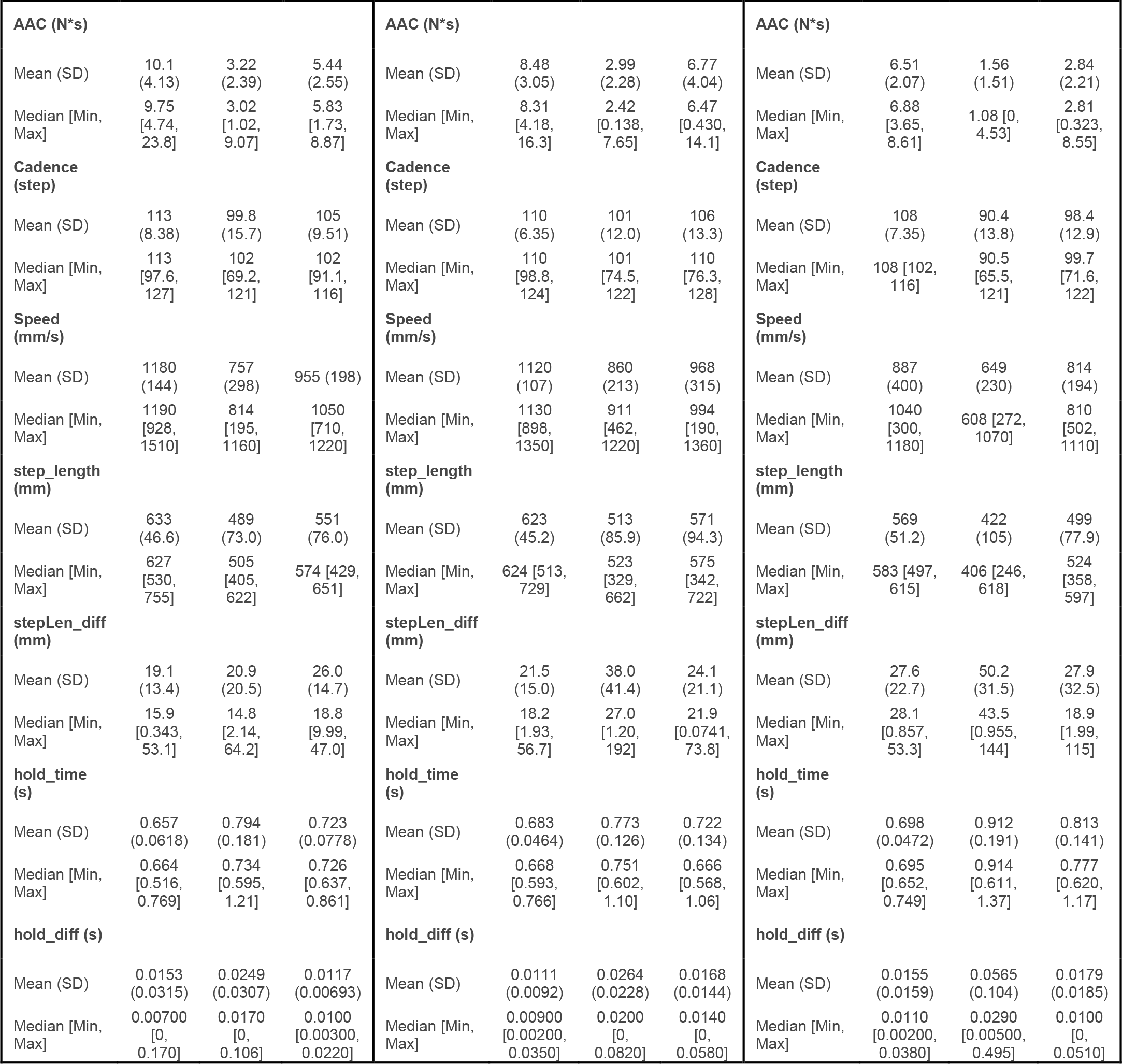
Detailed statistics of all kinematic features among all subjects, stratified by weight groups.

The normal-weight group (18.5 <= bmi < 25) consists of 30 healthy volunteers (HEA), 10 HOA patients (HOA), and 9 patients after the THA surgery (TRE). With a p-value of 6.5 * 10^−7^ from two-sample independent t-tests, AAC was the most significant feature distinguishing HEA and HOA. The HEA group had a mean AAC of 10.1 N*s (SD = 4.13), while the HOA group had a mean AAC of only 3.22 N*s (SD = 2.39). Speed, cadence, step length, and hold time were also significant features. For the TRE group, although a trend of shifting towards the HEA group was observed in AAC, step length, and hold time, t-test indicated that none of these shiftings was significant, and these variables in the TRE group still had significant differences between the HEA group.

The overweighted group (25 <= bmi < 30) consists of 25 HEA, 23 HOA, and 16 TRE subjects. AAC continues as the most significant feature distinguishing HEA and HOA. The HEA group had a mean AAC of 8.48 N*s (SD = 3.05), while the HOA group had mean AAC of 2.99 N*s (SD = 2.28), with a p-value of 7.5 * 10^−9^ from two-sample independent t-test. Speed, hold time, hold time difference, step length, and cadence also showed significant differences between HEA and HOA. The treatment significantly boosted the TRE group’s mean AAC to 6.68 N*s (SD = 4.42), making the difference between the HEA group non-significant. Cadence, speed, step length, hold time, and hold time difference showed a trend of shifting towards the HEA group, while none of them shifted significantly as AAC.

The obese group (bmi > 30) consists of 4 HEA, 21 HOA, and 19 TRE subjects. Hold time became the most significant feature with a p-value of 2.2*10^−4^. The HEA group has a mean hold time of 0.698 seconds (SD = 0.0472), and the HOA group has a mean hold time of up to 0.912 seconds (SD = 0.191). AAC, cadence, and step length are also significant features. The treatment significantly shifted the HOA subjects’ AAC, speed, step length, and step length difference towards the healthy subjects.

The increase in the ratio of HOA subjects as weight group increases infers BMI as a significant feature distinguishing HEA and HOA subjects. Figure 5 below plots the BMI among subjects with available demographic and diagnostic data. The 62 HEA subjects have a mean BMI of 24.9 (SD = 3.52), and the 61 HOA subjects have a mean BMI of 29.4 (SD = 5.46). Two-sample independent t-test shows a p-value of 5.3*10-7.

**Figure 5.**
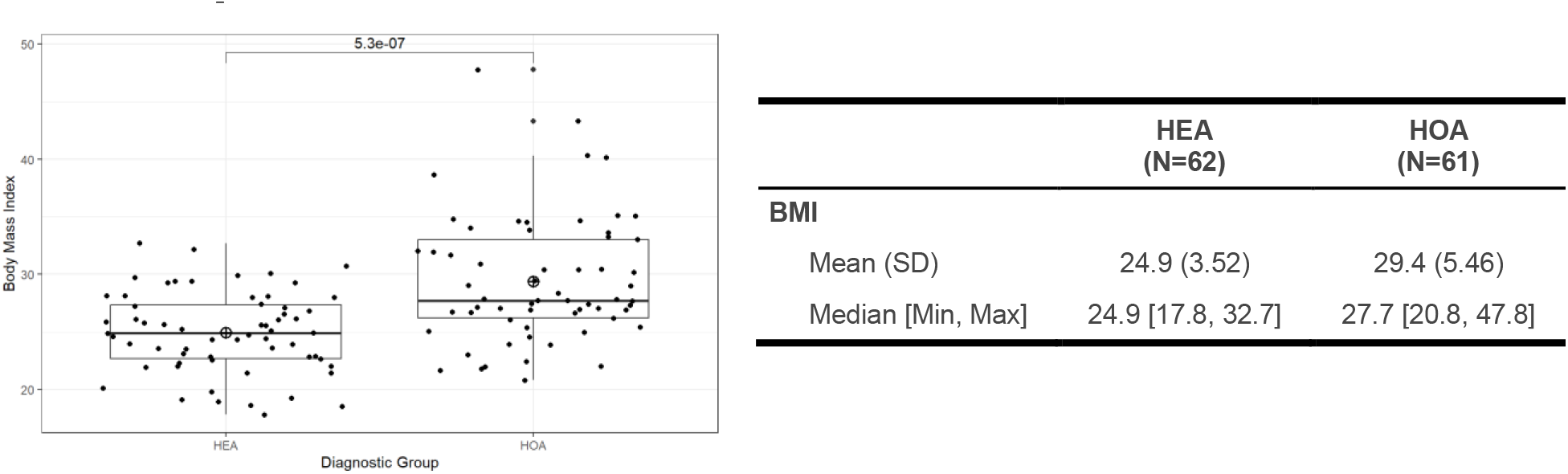
BMI among subjects with available demographic and diagnostic data.

Figure 6 below shows the accumulated GRF curve of all subjects, stratified by weight group. A subject from HOA or TRE tends to generate a curve with two high peaks and a deep smooth concave between them. While a subject from HOA tends to generate a curve with a longer horizontal span, lower peaks, and a shallower, less smooth concave in between. The THA treatment not only boosted the AAC of the TRE group but also help to reshape the GRF curves back to the general pattern of the HEA group.

**Figure 6.**
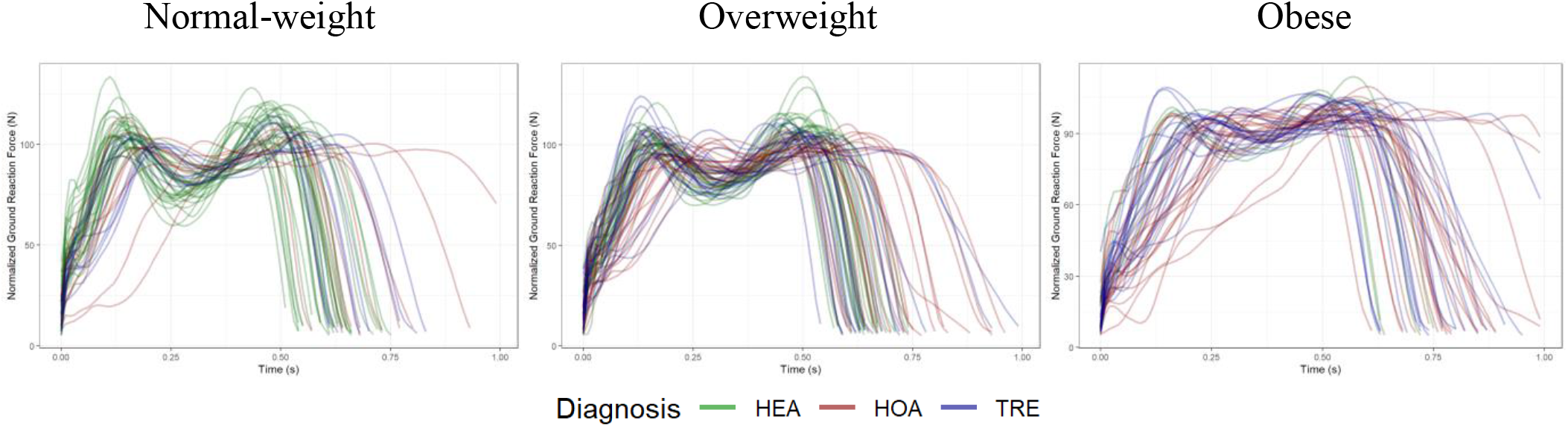
Normalized GRF curves generated by HEA, HOA, and TRE subjects, stratified by weight groups.

**Figure 7.**
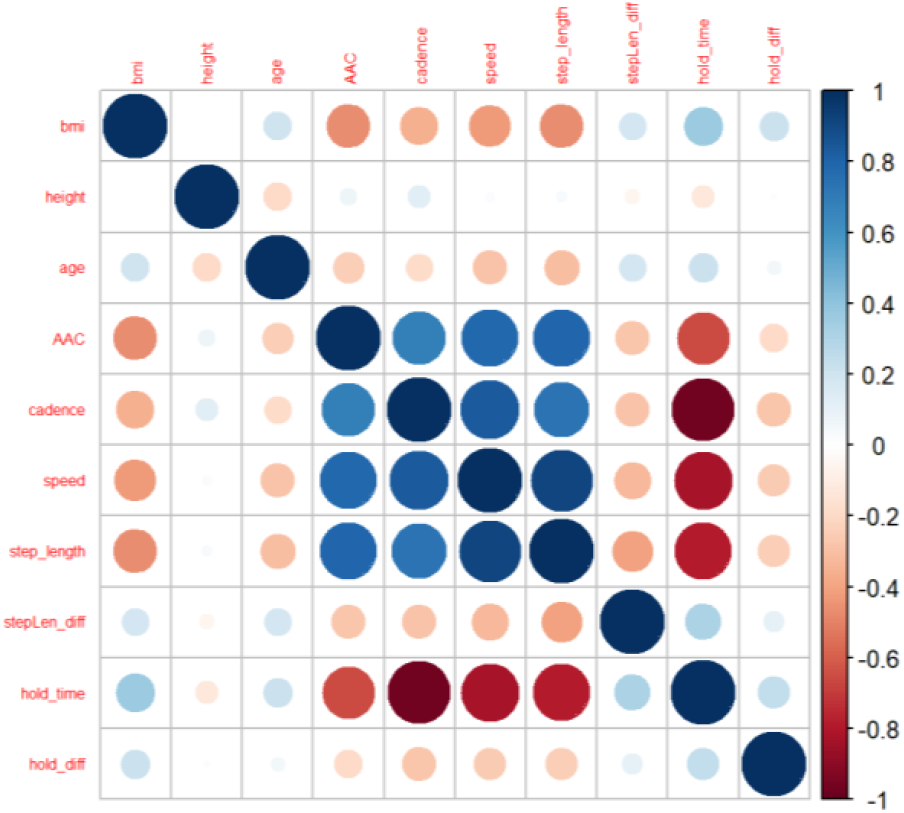
Correlation matrix of demographic and gait kinematic variables.

**Figure 8.**
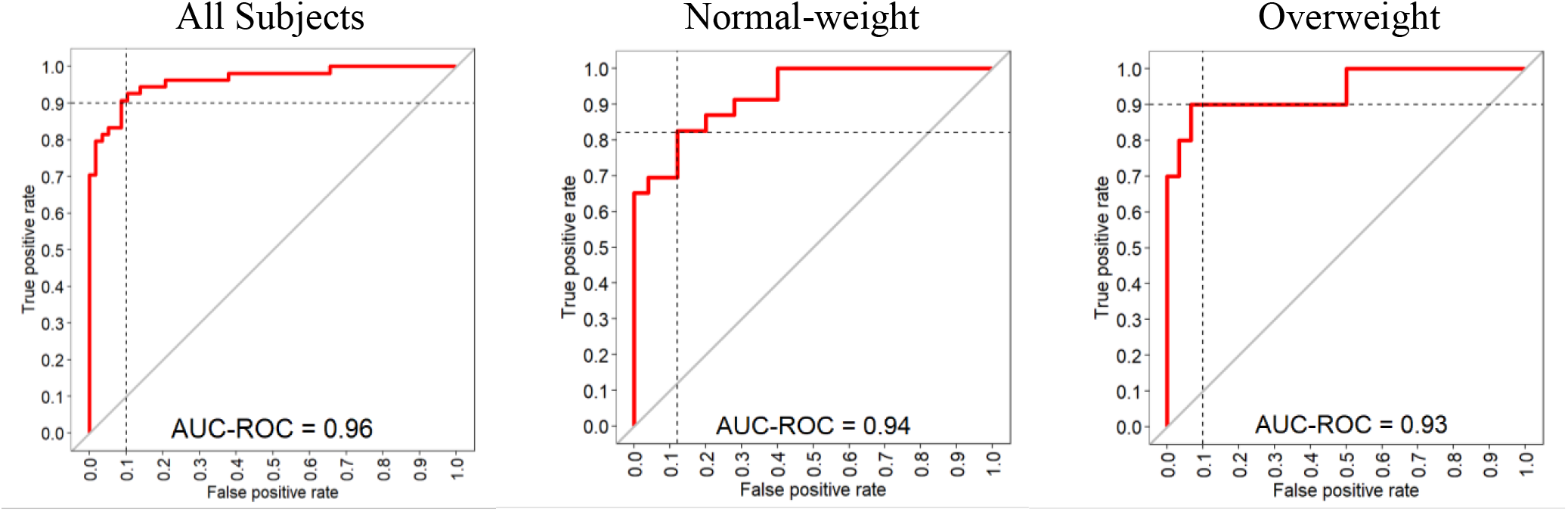
ROC curves of the penalized logistic HEA/HOA classifier on normal-weight, overweight, and all subjects.

It is worth noting that inner correlations exist among the gait kinematic variables. There is a pairwise positive correlation between AAC, cadence, speed, and step length. And all four of them have negative correlations with hold time. These correlations are in sync with common sense: a faster subject would walk with longer step length, strike more steps per minute and spend less time holding each foot on the ground. These variables’ correlations to AAC indicate that AAC could also be considered a measurement of walking dynamics.

Penalized Logistic Regression was trained as a binary classifier to diagnose HOA patients from the study population. The model was fitted on normal-weight subjects, overweight subjects, and all available subjects. No independent model was fitted on the obese group because of the severe data unbalancing. The results from repeated 5-fold cross-validation showed: on normal-weight subjects, the classifier achieved >80% sensitivity under 87% specificity, with an AUC of 0.94. On overweight subjects, the classifier achieved 90% sensitivity under 90% specificity, with an AUC of 0.93. And finally, on all subjects, the final classifier was able to achieve > 90% sensitivity under 90% specificity, with an AUC up to 0.96. These performance results indicate that penalized logistic regression could be a reliable tool to diagnose HOA patients from healthy subjects in a population.

Penalized logistic regression incorporates a penalty term to the model to address overfitting, which occurs when the model is over-complex with too many variables [6]. The penalty term shrinks the coefficients of less important variables towards zero, thereby regularizing the model and improving its performance [6]. Table 2 shows the coefficients and variable importance of the final model incorporating all subjects. As expected from inner correlations, coefficients of cadence, speed, and hold time shrunk to zero. The model chose hold difference as the most important predictor, followed by AAC and BMI. Interpreting the coefficients, we have:

♦ 1 second increase in hold difference multiplies the odds of HOA by 870. (Average hold difference of healthy subjects is only around 0.01 seconds)
♦ 1 N*s increase in AAC is associated with a 46.4% decrease in the odds of HOA.
♦ 1 unit increase in bmi is associated with a 10.6% increase in the odds of HOA.
♦ 1 year increase in age is associated with a 1.6% increase in the odds of HOA.
♦ 1 mm increase in step length is associated with a 1% decrease in the odds of HOA.
♦ 1 mm increase in step length difference is associated with a 0.27% increase in the odds of HOA.

**Table 2.** Coefficients and variable importance of the final penalized logistic model on all subjects.

| Variables | Coefficient | Importance(%) |
| --- | --- | --- |
| (Intercept) | 5.224 | NA |
| hold_diff | 6.769 | 100.0 |
| AAC | -0.624 | 9.218 |
| bmi | 0.101 | 1.498 |
| age | 0.016 | 0.238 |
| Step length | -0.0100 | 0.148 |
| stepLen_diff | 0.0027 | 0.0403 |
| cadence | 0.0 | 0.0 |
| speed | 0.0 | 0.0 |
| hold_time | 0.0 | 0.0 |

## V. Conclusions & Discussion

Clinical gait measurement is widely utilized in occupational therapy to quantify gait kinematic impairments resulting from diseases such as OA. Dr. Bertaux’s team at the University of Bourgogne Franche-Comté conducted such gait measurements on healthy volunteers and unilateral hip OA patients using force-sensing plates and motion capture systems [3]. The same measurements were taken on some of the patients six months following total hip arthroplasty treatments [3]. Based on Bertaux et al.’s public dataset, stratified by BMI weight groups, we built a computational pipeline to identify seven kinematic features that significantly differentiate HOA patients from healthy individuals. These features, along with demographic variables such as BMI, took part in penalized logistic regression models for HOA diagnosis with interpretable predictors. Cross-validation results showed that the final diagnostic model achieved over 90% sensitivity with 90% specificity and an AUC of 0.96. For the evaluation of THA treatment, several kinematic features of the THA-treated patients shifted significantly towards those of healthy volunteers. The treated patients’ ground reaction curves were also reshaped back to the general pattern of the HEA group. While differences between the treated and healthy groups still persisted.

Among the computed kinematic features, our proposed Ground Reaction Force Area-Above-Curve (AAC) proved to be a credible feature distinguishing HEA and HOA. T-test of AAC between HEA and HOA resulted in the lowest p-value on both normal-weight and overweight groups. AAC was also ranked as the second most important variable in the final penalized logistic regression model. While the exact interpretation of AAC still requires further research. Physically speaking, the area under the GRF curve is the total force impulse in Newton-second generated by the foot to the ground in each step [8]. Since AAC represents the concaved area formed by the first two GRF local maximum points, AAC could be interpreted as a measurement for the total variation of impulse between the two maximums. AAC’s positive correlations between general kinematic variables and the negative correlation between hold time indicate its ability to quantify walking dynamics.

It is worth noting that all kinematic variables in our pipelines can be collected without a motion capture system. In occupational therapy, there is a highly standardized technique to measure general gait kinematic variables (such as step length, hold time, and speed) using specialized rulers on the ground. The hold time of each step could also be coded from the video recordings. The only new equipment needed is the ground reaction force plate, which usually costs no more than $350. A simple and cheap diagnostic model could be easily applied to the general population for vast diagnosis. While a motion capture system would also excavate more significant kinematic features from the upper body, such as body balance, the 3D motion of the center of mass, etc. Further exploration of Bertaux et al.’s dataset is needed to build up consensus on HOA kinematic variables, and eventually, achieve a high-performance and cost-balanced HOA diagnostic model.

## Data Availability

All data produced are available online at: https://www.nature.com/articles/s41597-022-01483-3

## VII. Acknowledgement

We would like to acknowledge Dr. Katherine Dimitropoulou (Columbia University Programs in Occupational Therapy) for her suggestions during the early stage of this analysis.

## References

1. Thompson, Dave. “Variables of Stride Analysis.” Stride Analysis, University of Oklahoma Health Sciences Center, 24 Apr. 2002, https://ouhsc.edu/bserdac/dthompso/web/gait/knmatics/stride.htm#.

2. Bahl, J.S., et al. “Biomechanical Changes and Recovery of Gait Function after Total Hip Arthroplasty for Osteoarthritis: A Systematic Review and Meta-Analysis.” Osteoarthritis and Cartilage, vol. 26, no. 7, 20 Feb. 2018, pp. 847–863., https://doi.org/10.1016/j.joca.2018.02.897.

3. Bertaux, Aurélie, et al. “Gait Analysis Dataset of Healthy Volunteers and Patients before and 6 Months after Total Hip Arthroplasty.” Scientific Data, vol. 9, no. 1, 2022, https://doi.org/10.1038/s41597-022-01483-3.

4. Costello, K.E., et al. “Ground Reaction Force Patterns in Knees with and without Radiographic Osteoarthritis and Pain: Descriptive Analyses of a Large Cohort (the Multicenter Osteoarthritis Study).” Osteoarthritis and Cartilage, vol. 29, no. 8, 20 Mar. 2021, pp. 1138–1146., https://doi.org/10.1016/j.joca.2021.03.009.

5. Fang, Xin, et al. “Reference Values of Gait Using APDM Movement Monitoring Inertial Sensor System.” Royal Society Open Science, vol. 5, no. 1, 2018, p. 170818., https://doi.org/10.1098/rsos.170818.

6. Kassambara, et al. “Penalized Logistic Regression Essentials in R: Ridge, Lasso and Elastic Net.” STHDA, Statistical Tools for High-Throughput Data Analysis, 11 Mar. 2018, http://www.sthda.com/english/articles/36-classification-methods-essentials/149-penalized-logistic-regression-essentials-in-r-ridge-lasso-and-elastic-net/.

7. “Niams Health Information on Osteoarthritis.” National Institute of Arthritis and Musculoskeletal and Skin Diseases, U.S. Department of Health and Human Services, 8 June 2022, https://www.niams.nih.gov/health-topics/osteoarthritis.

8. Wiik, Anatole Vilhelm, et al. “Abnormal Ground Reaction Forces Lead to a General Decline in Gait Speed in Knee Osteoarthritis Patients.” World Journal of Orthopedics, vol. 8, no. 4, 18 Apr. 2017, p. 322., https://doi.org/10.5312/wjo.v8.i4.322.

